# Atlas of epistasis

**DOI:** 10.1101/2021.03.17.21253794

**Authors:** Clément Chatelain, Samuel Lessard, Vincent Thuillier, Cedric Carliez, Deepak Rajpal, Franck Augé

**Affiliations:** SANOFI R&D, Translational Sciences, Chilly Mazarin, 91385, France; SANOFI R&D, Translational Sciences, Framingham, MA 01701, USA; SANOFI R&D, Data & Data Sciences, Chilly Mazarin, 91385, France

## Abstract

We performed a genome-wide epistasis search across 502 phenotypes in case control matched cohorts from the UK Biobank. We identified 152,519 genome wide significant interactions in 68 distinct phenotypes, and 3,398 interactions in 19 phenotypes were successfully replicated in independent cohorts from the Finngen consortium. Most interactions (79%) involved variants that did not present significant marginal association and might explain part of the missing heritability for these diseases. In 10 phenotypes we show the presence of epistasis between common variants with intermediate to large effect size (*OR* > 2) supporting the hypothesis that common diseases are modulated by common variants. Most of the variants in interactions (82%) were more than 1Mb apart and cis-epistasis was hardly found outside the HLA region. Functional annotation of the variants suggests that most mechanisms behind epistasis occurs at the supra pathway level and that intra-gene or intra-pathway epistasis is rare. Surprisingly we find a significant biais toward antagonistic epistasis, representing 60% to 95% of interactions. In type 1 diabetes, hypothyroidism, disorders of mineral absorption, rheumatoid arthritis, asthma, and multiple sclerosis more than 50% of interactions were completely compensating the effect of the marginally associated variant. In psoriasis we identified an interaction between a stop gain variant in CCHCR1 with two missense variants in MUC22 and HSPA1L leading to a 3 fold increase of the effect of CCHCR1 variant on disease risk. Our study shows that there is still much to discover in epistasis and we provide the full summary statistics results to researchers interested in studying epistasis.

## Introduction

The emergence in the recent years of human genetic data from large-scale biobanks, such as UK Biobank, Finngen, and Biobank Japan ^1–3^, has been followed by a new wave of Genome Wide Association Studies (GWAS) spanning the entire human phenome ^4–7^, while investigating variation across populations ^6,7^. These studies provided new systematic estimate of narrow sense heritability ^4,8^, the proportion of phenotypic variation due to additive genetic variation. More importantly the summary statistics resulting from these efforts represent now a key resource beside GWAS catalogs ^9^ for many researchers to explore the genetic architecture of human diseases ^10^.

However such PheWAS rarely consider potential nonlinear relationship between genotype and phenotypes. In particular interactions between genetic variants, a phenomenon known as epistasis, are not considered. Epistasis is generally defined as deviation from additivity of the combined effect of multiple variants and is suspected to be an ubiquitous property of genetic architecture ^11^. Molecular causes of epistasis may range from intra-protein interactions ^12^, transcription regulation ^13^, to molecular recognition, substrate competition, functional redundancy within pathways, or higher level interactions between pathways ^14^. Epistasis has long been identified as a potential explanation for the discrepancy observed between narrow and broad sense heritability, i.e. the proportion of phenotypic variation due to total genetic variation ^15,16^. Estimations by Zuk et al. suggests that epistasis could be responsible for up to 80% of such missing heritability ^16^. More recent estimates by Young et al. ^17^ still present a gap of 33%. Finally epistasis has been shown to be a key component of evolution and might have important application in therapeutic target identification and population stratification.

In the past decade a number of statistical approaches have been developed to detect epistasis, ranging from regression ^18^ and linear mixed models ^19^ to machine learning ^20^ approaches, and are reviewed elsewhere ^21,22^. However, despite the suspected importance of epistasis in the genetic architecture of the human phenotypes, the application of these methods to the systematic study of epistasis has mostly remained limited to a small number of illustrative phenotypes, almost exclusively among the 7 diseases of the WTCCC ^23^. Wang et al. ^18^ performed one of the first attempts of genome wide search for epistasis on the WTCCC cohorts and identified significant interactions in Type 1 Diabetes (T1D) and Crohn’s Disease (CD), almost all localized within MHC region. Subsequent genome wide epistasis studies ^19,24^ of these cohorts identified additional interactions in Rheumatoid Arthritis (RA), Coronary Artery Disease (CAD), Bipolar Disorder (BD), Type 2 Diabetes (T2D) and Hypertension (HT), again mostly localized within MHC region.

Multiple obstacles explain why genome-wide epistasis studies have remained so limited. Epistasis effect size has been suspected to be small and therefore to require unrealistic sample size to be detected ^25^. Most epistasis detection methods do not implement correction for confounding factors in particular to account for population structure. Testing all genotyped variant pairs for epistasis remains a computationally challenging task, in particular for those methods correcting for confounding factors such as FastLMM ^19^, which required 950 compute years to analyze 7 diseases. Additional obstacles rise when going at the biobank scale. The presence of relatives, complex population structure, and strong case control imbalance makes accurate control of type error rate more challenging. In univariate GWAS analysis, dedicated solutions based on linear mixed models such as SAIGE ^5^ have been developed to address this point but do not yet implement epistasis tests. Finally the required computational resource increases as the number of phenotypes considered in the biobank and linear mixed models and even logistic regression with covariates cannot be realistically applied.

Here we present a genome wide epistasis analysis on 502 diseases from the UK Biobank with more than 500 cases using the likelihood ratio test introduced in BOOST ^18^. In a previous study we show that BOOST performs accurate type 1 error rate control even in presence of linkage disequilibrium while reaching satisfying statistical power compared to other tools ^26^. In order to control for confounding factors, we prepare case control matched cohorts for each phenotype following the method described by Luca et al.^27^. Significant interactions are replicated in the Finngen biobank and discussed thereafter.

## Materials and methods

### Ethics statement

#### Participants and data sources

The UK Biobank is a population-based prospective cohort composed of approximately 500,000 individuals across the United Kingdom combining genotype and deep phenotyping data. More details regarding the UK Biobank project have been published by Bycroft et al. ^1^.

Finngen is a public–private partnership project combining genotype data from biobanks and electronic health record data from Finnish public registries. It aims at sequencing 500,000 Finnish individuals by 2023. In this study we use data from Finngen release 6 which includes genotype and phenotype data for 269,077 participants.

### Genotyping and quality control

UK Biobank participants genotypes were obtained from two genotyping arrays, the Affymetrix UK BiLEVE Axiom or Affymetrix UK Biobank Axiom array. For the purpose of this study we used a subset of 266,679 individuals satisfying all following quality control criteria: (i) identified as inlier in heterozygosity and missing rates (Data-field 22027), (ii) not identified as putatively carrying sex chromosome configurations other than XX or XY (Data-field 22019), (iii) selected as input for the phasing of autosomal chromosomes (Data-field 22028), (iv) without genetic kinship to other participant (Data-field 22021), (v) identified as white British and very similar genetic ancestry based on a principal components analysis of the genotypes (Data-field 22006). We restricted the analysis to SNPs from HRC imputation with minor allele frequency greater than 0.01, in Hardy Weinberg equilibrium (pHWE > 1e-10) and with call rate larger than 0.95. Details about quality control has been published by the UK Biobank ^1^. To insure the feasibility of an exhaustive bivariate epistasis analysis we restricted the study to the 576,429 genotyped variants passing the quality control criteria.

Finngen participants genotypes were obtained from Illumina and Affymetrix arrays and was composed of 52 datasets. A total of 16,962,023 variants were imputed using 3,775 whole genomes from the Finish specific reference panel SISu v3 (*http://sisuproject.fi/*). We used Finngen as replication cohort including only variants presenting significant epistasis in the discovery phase with the UK Biobank. In order to limit the population structure we considered 182,584 unrelated individuals of Finish ancestry satisfying the following quality control criteria: (i) non-ambiguous gender, (ii) genotype missing rate lower than 5%, (iii) no excess heterozygosity (*±4SD*), (iv) of finish ancestry, defined by PCA of Finngen samples together with EUR and FIN samples from the 1000 genomes projects, (v) no related individual up to degree 2 as computed from kinship matrix.

### Phenotypes and cohort selection

#### UK Biobank

In the UK Biobank data we derived the phenotypes from primary and secondary hospital diagnose data (Data-field 41270) using ICD10 codes of level 3. An individual was defined as a case for a particular ICD10 code (eg. J45 asthma) if it had at least one registered diagnose for this ICD10 code or any subcode (eg. J45.8 mixed asthma). The age of first diagnose was defined as the earliest diagnose of these codes (Data-Field 41280). The current age of the individual was defined as his age at the latest date of entry of all hospital diagnose in the dataset, March 2017.

A major difficulty in performing an epistasis scan across the entire genome for a large number of phenotypes is the computational cost. Using linear mixed method approaches such as SAIGE ^5^ would be computationally unfeasible. Even logistic regression models for epistasis including continuous covariates would be too computationally prohibitive.

First, we divided the cohort into genetic clusters without significant population structure with the following process. We performed a principal component analysis (PCA) on all selected individuals using 100,000 pruned variants listed by UK Biobank. The 10 first components presented significant population structure (Tracy Widom test pval<1e-2, Table S4) and were selected to perform hierarchical clustering of all individuals in the cohort using Ward’s algorithm (Figure S1). We evaluated iteratively the presence of population structure in each cluster starting from the hierarchy root cluster by performing a PCA and a Tracy Widom test. In absence of significant population structure (Tracy Widom pval>1e-2 for the PCA first eigenvalue) we selected the cluster, otherwise we tested the two subclusters. We removed clusters with 20 individuals or less excluding 787 individuals. Outlier removal (*±6 SD*on the first 5 PCs) for each cluster excluded 41 additional individuals. The remaining 265,538 individuals were distributed into 276 clusters, with clusters of more than 1,000 individuals accounting for a large majority of individuals (225,099, Table S5 and Figure S2). We used the FLASHPCA2 software for the PCAs ^28^, the tw function in the EIGENSOFT suite to perform Tracy Widom test ^29^, and the fastcluster R library to perform hierarchical clustering ^30^.

For a given phenotype we matched each case to *n* controls uniformly sampled from controls of the same sex, same genetic cluster and older than the case at the time of its first diagnosis (Figures S4 and S5). We optimized the control case ratio *n* for each phenotype to maximize the effective sample size 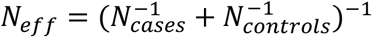. An example of optimization is depicted in Figure S3.We excluded phenotypes with less than 500 cases and included 502 remaining phenotypes in the study.

We performed simulations for each phenotype to estimate the statical power with the following parameters: phenotype model with only epistatic term *logit[E(D*|*X*_1_ *= x*_1_ *X*_2_ *= x*_2_*)] = α+β*_12_*x*_1_*x*_2_ with *X*_1_ and *X*_2_ coding the genotype of two variants of minor allele frequency 0.1 and no linkage disequilibrium, number of cases and controls as identified by the previous optimization step, disease prevalence *K*_0_ as observed in the full UK Biobank cohort, pvalue threshold for genome-wide significance of 8e-12. We estimated the minimal epistasis odd ratio *OR* = exp[*β*_12_]detectable with statistical power 0.8 by root search with 100 replicates for each value of *β*_12_. All estimates are reported in Table S1 and ranged from OR=1.07 for hypertension (I10) to OR=2.9 for pancreas cancer (C29).

#### Finngen

In the Finngen data, used here for replication, about 2700 phenotypes have been manually defined by Finngen expert groups from health registry data ^2^. We manually mapped to Finngen phenotypes the 102 phenotypes selected in the UK Biobank and presenting at least one significant epistatic interaction (mapping provided in Table S1).

We applied the previous case control matching process to define Finngen replication cohorts. A total of 134,080 individuals were selected and distributed across 3008 genetic clusters.

### Epistasis analysis

#### Discovery

We performed exhaustive bivariate epistasis tests on each UK Biobank phenotype using BOOST epistasis testing implemented in plink 1.9 with the options ‘–fast-epistasis boost –epi1 0.001’ which discarded results with epistasis pvalue higher than 1e-3, to keep results files of reasonable size (∼5Gb per phenotype). The genome-wide epistasis scan of the 502 phenotypes required 2880 cpus during approximately 7 days. For each phenotype we computed the false discovery rate with the Benjamini and Hochberg method ^31^ setting to 1 the pvalue of tested but discarded variant pairs. We considered variant pairs with false discovery rate below 0.25 as presenting significant epistasis. The genomic inflation factor at P=0.001 was lower than 1.1 in all phenotypes and lower than 1.05 in 94% of phenotypes showing a good control of type 1 error rate (Figure S7 and S8).

#### Clumping

We clumped significant results from the discovery study by iteratively selecting lead variant pairs (*x*_1_ *x*_2_) with lowest epistasis pvalue and excluding all variant pairs (*x′*_1_, *x′*_2_) within 250kb with 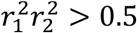 where 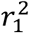 (resp. *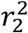*) is the Linkage Disequilibrium (LD) between *x*_1_ and *x′*_1_ (resp. *x*_2_ and *x*′_2_). We excluded lead variant pairs with strong LD (*r*^2^ > 0.9).

#### Univariate and bivariate models

For each phenotype we first tested the marginal association of each variant involved in epistasis with a simple Fisher test using plink 1.9 option ‘–assoc fisher’ without covariate on our case control matched cohort. We then obtained a more powerful and complementary evaluation of the marginal association from the population meta-analysis study published by the Pan-UKBB team ^6^. To evaluate if a variant would be detected by a genome-wide univariate analysis we used a a Bonferonni corrected pvalue threshold of 8.67e-8. Finally we fitted a bivariate logistic regression model on each variant pairs *X*_1_ and *X*_2_ with significant epistasis: logit[*E*(*D*|*X*_1_ = *x*_1_, *X*_2_ = *x*_2_)] = *α* + *β*_1_*x*_1_ + *β*_2_*x*_2_ + *β*_12_*x*_1_*x*_2_. The modulation of the effect of variant *X*_1_ by variant *X*_2_ is derived from this model as (1 + *β*_12_/*β*_1_*x*_2_) with *β*_12_/*β*_1_ the epistatic factor.

#### Replication

We tested for epistasis all variant pairs presenting significant epistasis in a UK Biobank discovery cohort in the corresponding Finngen replication cohort using plink option ‘–fast-epistasis boost’. We considered as successful replicated interactions with false discovery rate below 0.05.

### Variant to gene and pathway mapping

We mapped variants to genes using three types of data: variant transcript consequence computed with VEP, eQTL mapping from open target genetics (see Ghoussaini et al. ^32^ for detailed eQTL sources), and pQTL mapping from Sun et al. study ^33^. We evaluated the presence of known physical interaction between genes using the protein protein interaction network from BioGRID Release 4.2.191. Finally we assigned genes to 2868 canonical pathway obtained from MSigDB Collections.

### Heritability and variance explained

To estimate disease heritability, we generated GWAS (univariate) summary statistics for each phenotype using plink2 (--glm) with the ‘cc-residualize’ modifier ^34^, adjusting for sex, age, and the top 10 first principal components. We included imputed variants but restricted to variants with MAF>0.01 and present in the HapMap 3 variant reference panel. We used LD score regression to calculate additive SNP heritability, using pre-compiled LD scores from 1000 Genomes European dataset^35^. The heritability estimates were converted to the liability scale using the disease prevalence in the UK Biobank dataset (i.e. assuming it is the same as population prevalence). We used Fisher exact test to calculate the enrichment of significant interactions for diseases with *h*^*2*^>0.1, restricting to phenotypes with N_eff_ > 5,000.

To estimate the gain in variance explained, we fitted logistic regression models with and without the interaction term for all significant variant pairs, and calculated Nagelkerke’s pseudo-R2. For each variant pairs, the difference in pseudo-R2 between the full model (additive + interaction) and additive-only model was used to estimate the proportion of variance explained by epistasis.

## Results

### Significant variant pairs

In the discovery cohorts we detected 152,519 independent genome-wide significant interactions (FDR<0.25) in 68 phenotypes. The following 8 phenotypes presented from 200 to more than 100,000 interactions: Intestinal malabsorption (K90) and disorder of mineral metabolism (E83), type 1 diabetes (E10), rheumatoid arthritis (M069), hypothyroidism (E03), psoriasis (L40), asthma (J45), multiple sclerosis (G35). Other phenotypes presented less than 40 interactions (table 1). In the simulations the minimal epistatic odd ratio detectable between common variants ranged from 1.07 for hypertension(I10) to 2.92 for pancreas cancer (C25) and were smaller than 2 for 251 phenotypes. While median minimal odd ratio was significantly higher in phenotypes with no significant interactions (1.8 vs 2.2) there was no significant correlation with the number of interactions (Table S1), suggesting that this number was reflecting the etiology of these diseases rather than higher statistical power.

**Table 1:**
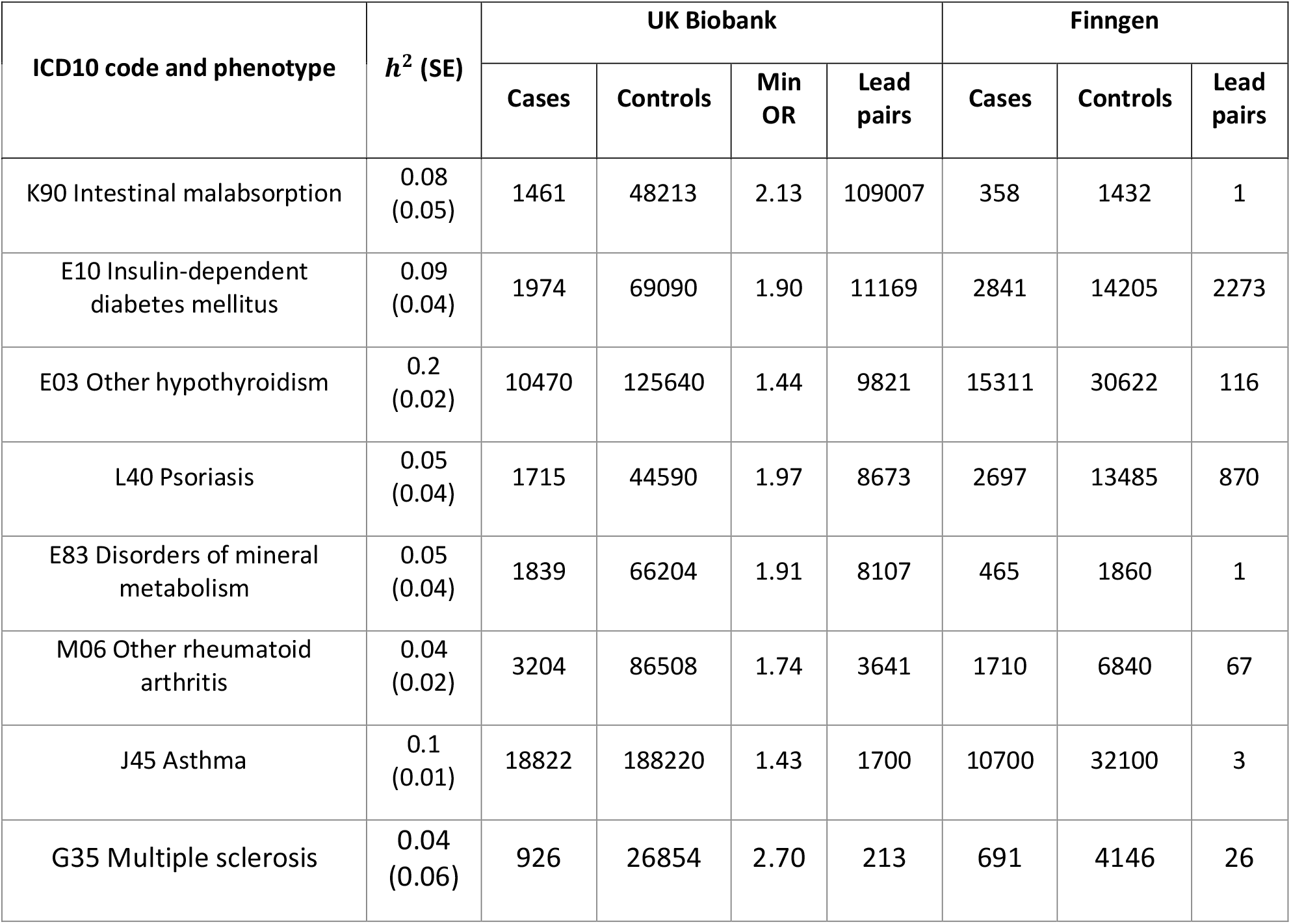
Number of independent variant pairs with significant epistasis in the UK Biobank discovery cohorts (FDR<0.25) and the Finngen replication cohorts (FDR<0.05), minimal epistasis odd ratio (OR = exp[β_12_]) detectable with power 0.8, and estimated additive heritability h^2^. Phenotypes with less than 40 significant interactions in discovery are reported Table S1.

In the replication cohorts 3,398 interactions in 19 phenotypes presented significant epistasis with a conservative threshold (FDR<0.05). An important proportion of interaction were reproduced for type 1 diabetes (20%), psoriasis (10%) and multiple sclerosis (12%). Only one interaction was reproduced for intestinal malabsorption and disorder of mineral metabolism, which might be explained by the limited number of cases in these replication cohorts, or the presence of an uncontrolled confounding factor for these phenotypes in the UK Biobank population.

### Enrichment in high impact region

#### Variant locations

A majority of variants involved in epistasis were located in intergenic (40%) and intronic regions (48%). However intergenic regions were significantly depleted in interacting variants (Fisher’s exact test pval<1e-173) while coding and intronic regions were significantly enriched (pval<1e-6). The variant distribution across genomic regions, the enrichment in coding and intronic region, and depletion in intergenic regions was consistently reproduced in all 7 diseases with more than 1000 significant interactions (Figure 1).

**Figure 1:**
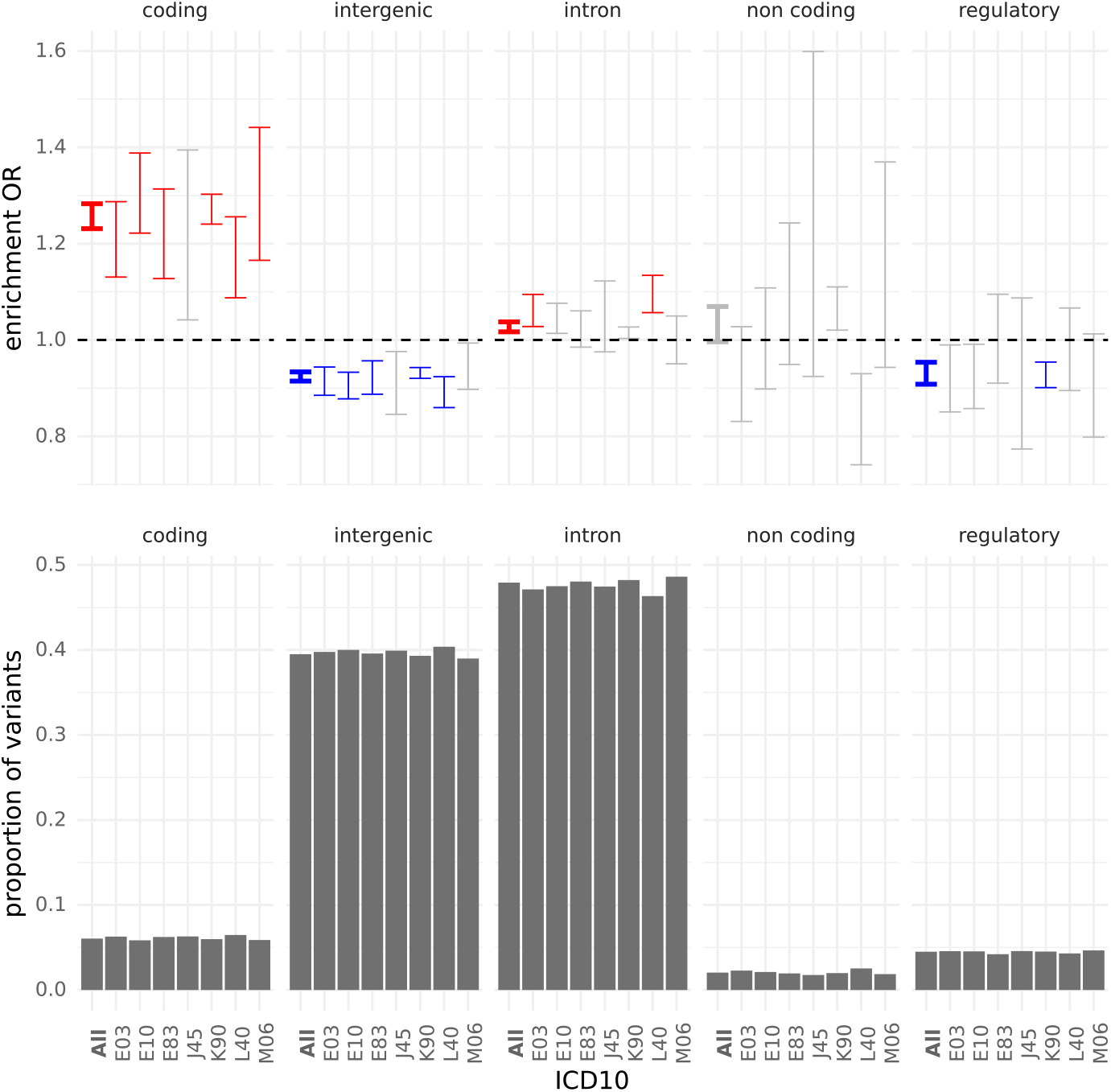
Distribution of variants with significant epistasis across 5 genomic regions categories defined from VEP annotation: regulatory region includes regulatory region and TF binding site variants, non coding includes mature miRNA and non coding transcript exon variants, intergenic includes intergenic, upstream and downstream gene variants. Results are given for the 7 diseases with more than 1000 interactions and for all diseases together (bold). Results are given for variant identified in the discovery cohorts. (top) Odd ratios (OR) estimates and 95% confidence intervals for enrichment in variants with significant epistasis compared to the number of variants sequenced in the region category. Blue error bars indicates significant depletion (Fisher’s exact test pval<1e-3) and red significant enrichment. (bottom) proportion of significant variant in each region.

#### Odd ratios for high impact variants

##### All identified

Among genotyped high impact variants (start loss, stop gain, stop loss, missense, splice donor and splice acceptor variants) 124 significant interactions were successfully replicated (table 2) for type 1 diabetes (E10) and psoriasis (L40), asthma (J45) and rheumatoid arthritis (M06). Bivariate and marginal contingency tables and odd ratios for each interaction are reported in Figure S11.

##### Loss of function

The interactions in psoriasis between the CCHCR1 stop gain variant (rs3130453) and two missense variants of MUC22 (rs3094672) and HSPA1L (rs2227956) are an example of synergistic epistasis (contingency tables reported in figure 2). While the stop gain variant is marginally associated to a weak increase in psoriasis risk (OR=1.49 pval=7.7e-20), the variant effect is increase by a factor 2.7 in the presence of two minor alleles of MUC22 missense variant (OR=3.2) and by a factor 3.1 in the presence of two minor alleles of HSPA1L missense. CCHCR1 is thought to be a regulator of mRNA metabolism and was already associated with psoriasis ^36^ but also with rheumathoid arthritis ^37^. MUC22 is coding for a protein of the mucin family present in epithelial secretions. HSPA1L is a gene located in the HLA region and coding for a heat shock protein with molecular chaperon function. The HSP gene family was previously associated with inflammation processes ^38^ and immune diseases (rheumatoid arthritis ^39^, multiple sclerosis ^40^, IBD ^41^, IPF ^42^, and COPD ^43^).

**Figure 2:**
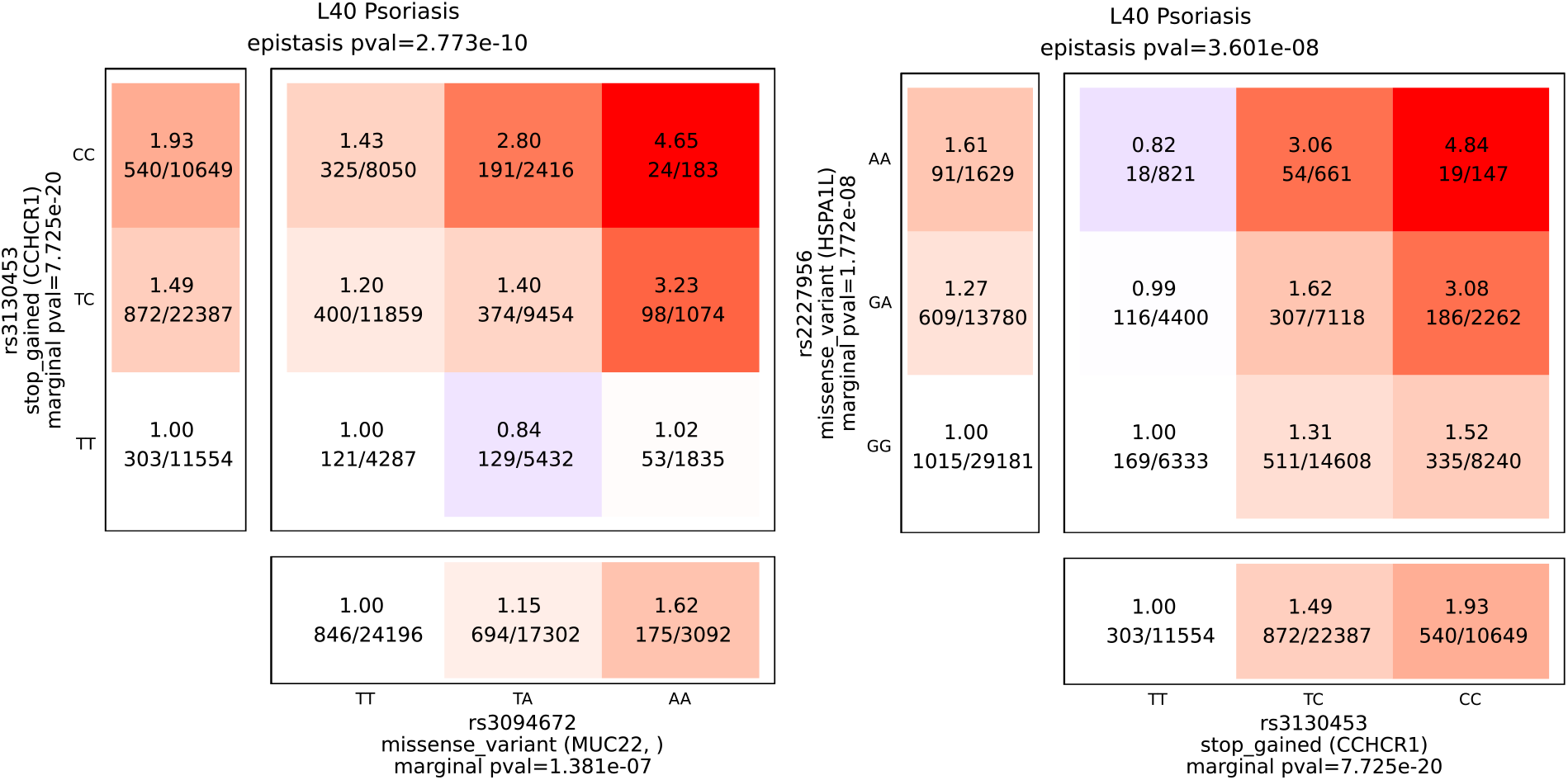
contingency tables for two interaction in psoriasis involving the CCHCR1 stop gain variant rs3130453. Marginal association pvalue were computed by Fisher’s exact test on the UK biobank psoriasis cohort. Data are represented for the UK Biobank discovery cohort.

### Distance between interacting variants

#### Chromosomes

After excluding the HLA region and taking into account the distribution of the sequenced variants on the chromosomes we observe a homogeneous distribution of the interactions across the chromosomes (Figure 3) with the exception of a high enrichment of interaction on chromosome 6 probably due to interactions within immune regulator regions. Most replicated interactions were intrachromosomal with a majority of interactions in the HLA region.

**Figure 3:**
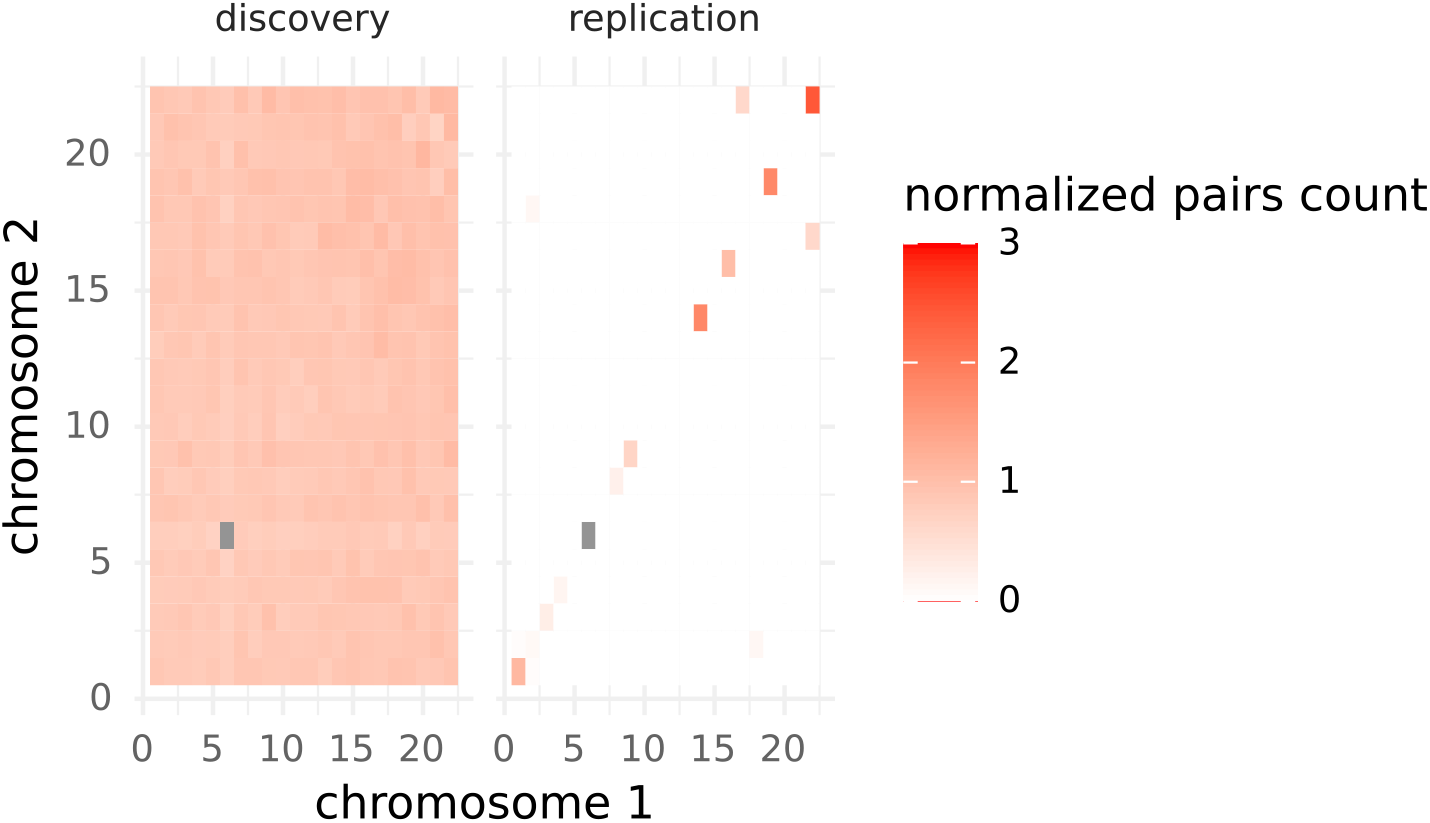
distribution of the intra and interchromosomal interactions normalized by the expected number of pairs for a uniform distribution, defined as 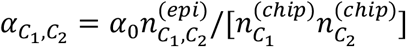 with 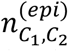 the number of variants pairs with significant epistasis between chromosomes C_1_ and C_2_, 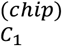 and 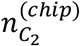 the number of sequenced variants in chromosomes C_1_ and C_2_, and a normalization factor 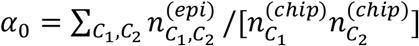 Enrichement on chromosome 6 was α =58 in the discovery cohorts and α = 473 in the replication cohorts.

#### Intrachromosome interactions

Intrachromosomal interactions are composed for 95% of variants distant from more than 1Mb and the distance distribution is very similar to the distribution of distance of uniformly distributed variants (Figure 4). The only exception is observed for 3 phenotypes (K90, J45, L40) for which we observe 60% of variant pairs distant from less than 1Mb, even after excluding HLA region.

**Figure 4:**
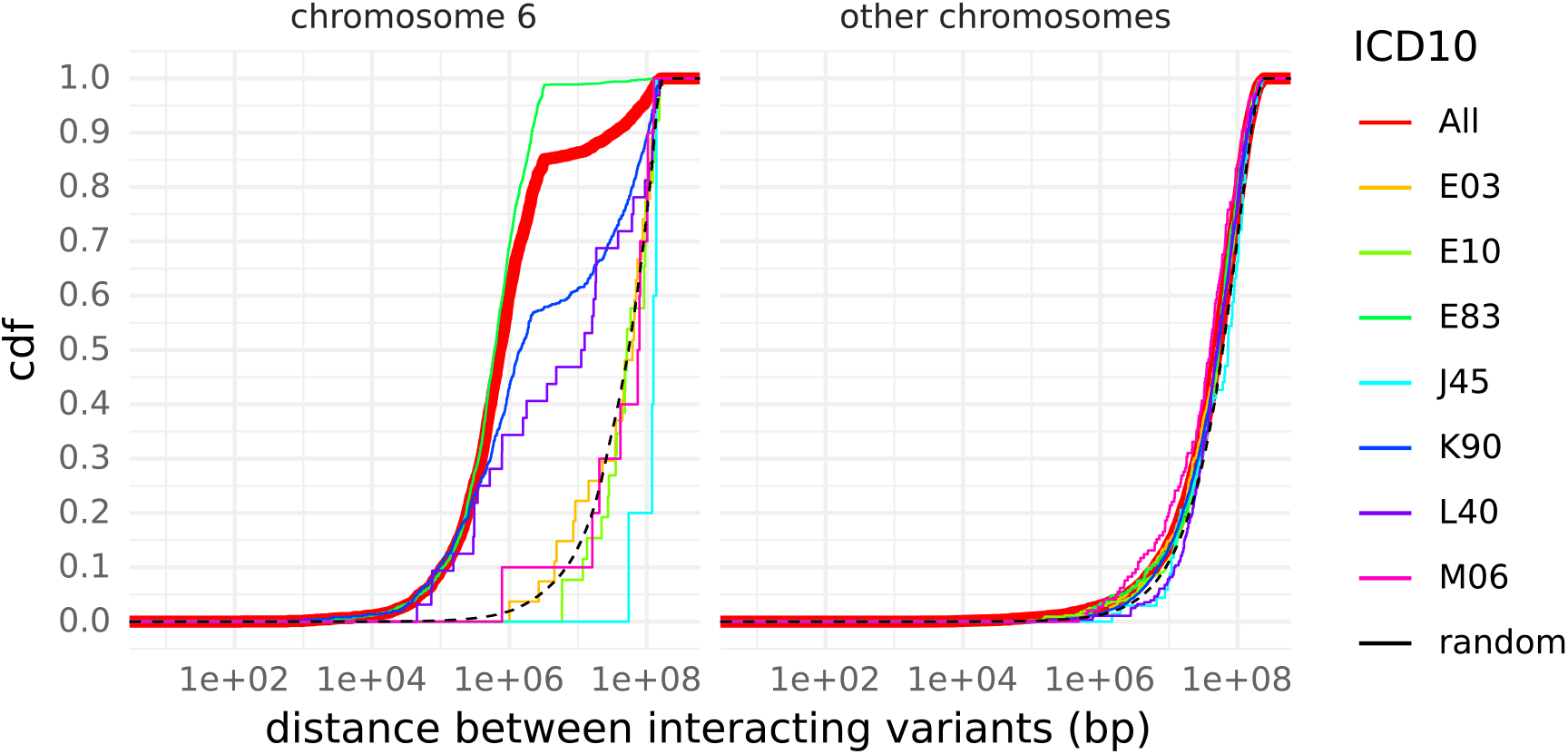
Distance distribution between interacting variants located on a same chromosome, on chromosome 6 (left) and on all other chromosomes (right). The distributions are given for the 7 diseases with more than 1000 interactions (colored thin lines) and for all diseases (thick red line). The dashed black line depicts the distance distribution in uniformaly distributed variants. Data is shown for the discovery cohort.

### Effect sizes

#### Comparison with univariate analysis and proportion of new discoveries

A majority of significant interactions occurred between variants that did not present significant marginal association in the present study (78%) or in the Pan-UKB study (79%). Only 15% of interactions occurred between 2 variants presenting both significant marginal associations (11% with both marginal association in the Pan-UKB study). We observed a similar distribution for all 7 diseases with largest number of interactions (Figure 5). This proportion was higher among the replicated interactions (24% and 36% for this study and the Pan-UKB study respectively).

**Figure 5:**
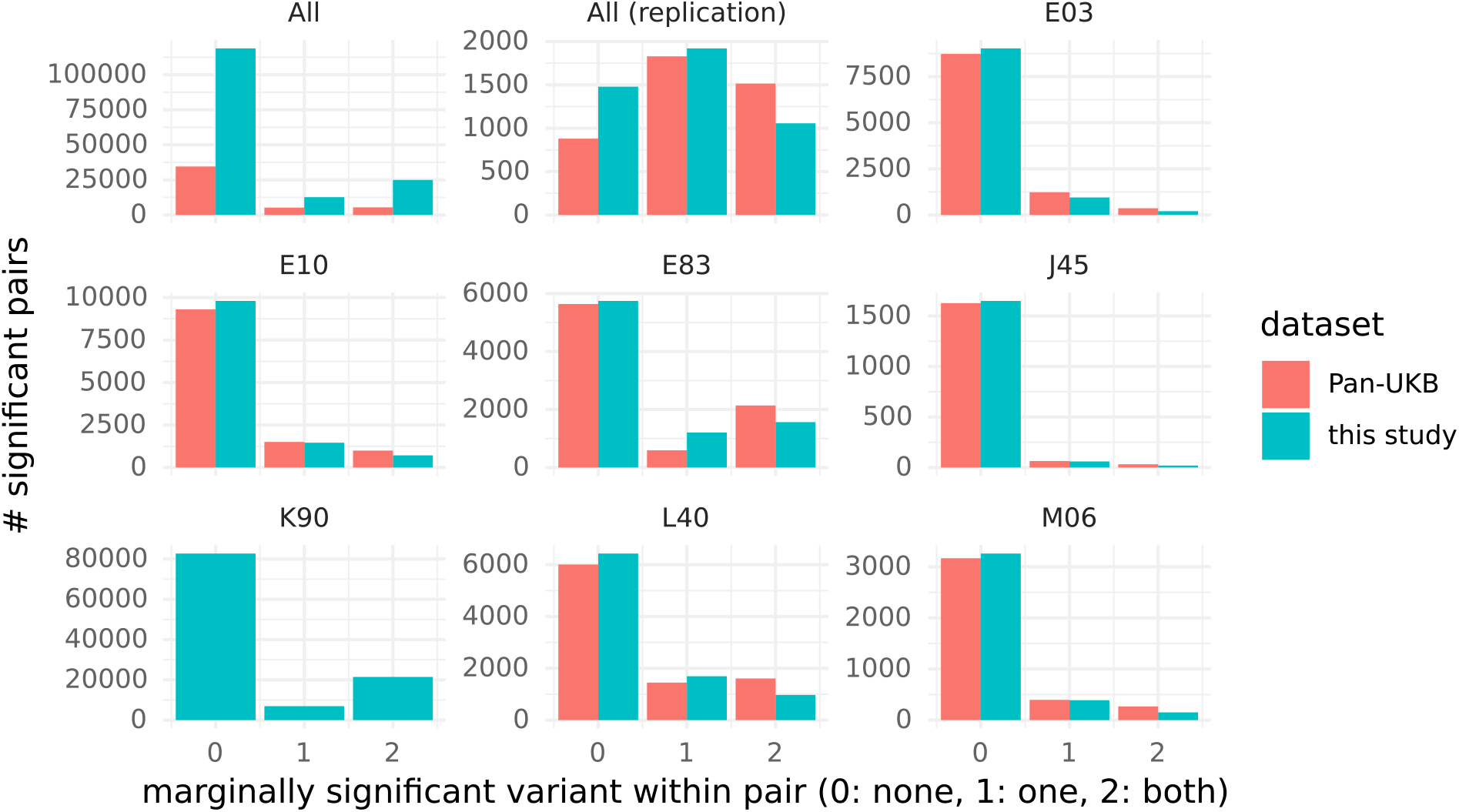
Number of variant pairs for which both variants do not present marginal association (0), only one variant present marginal association (1), or both variants present marginal association (2). Marginal association are tested in the present UK Biobank cohort with Fisher’s exact test (blue) and in the Pan UKB study ^6^ with SAIGE ^5^. The category ‘All’ includes all variants pairs significant in the discovery phase and the category ‘All replicated’ all variant pairs replicated.

#### Effect size of common variants

Among the significant interactions identified 3% presented intermediate effect size (*OR* = *exp*(*β*_12_) > 2 or < 0.5) and 0.1% presented high effect size (*OR* > 5 or < 0.2). Interestingly all the high effect size interactions were protective (Figure 6).

**Figure 6:**
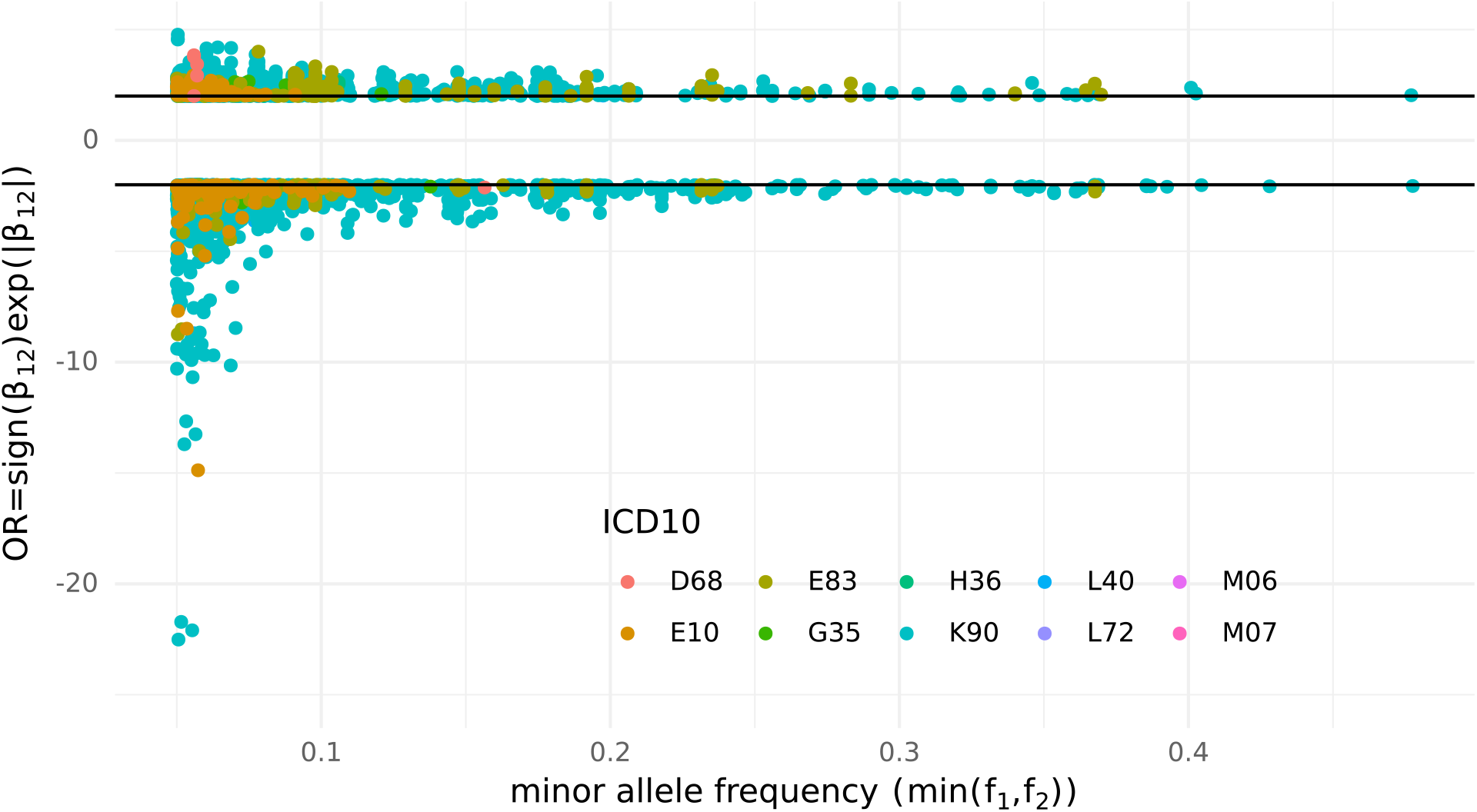
Effect size of the interaction estimated from the full bivariate logistic regression model coefficient β_12_, and effect allele frequency of variant 1 (f_1_) or variant 2 (f_2_) whichever is smaller. Only interactions with effect size OR > 2 or OR < 1/2 are represented.

**Figure 6:**
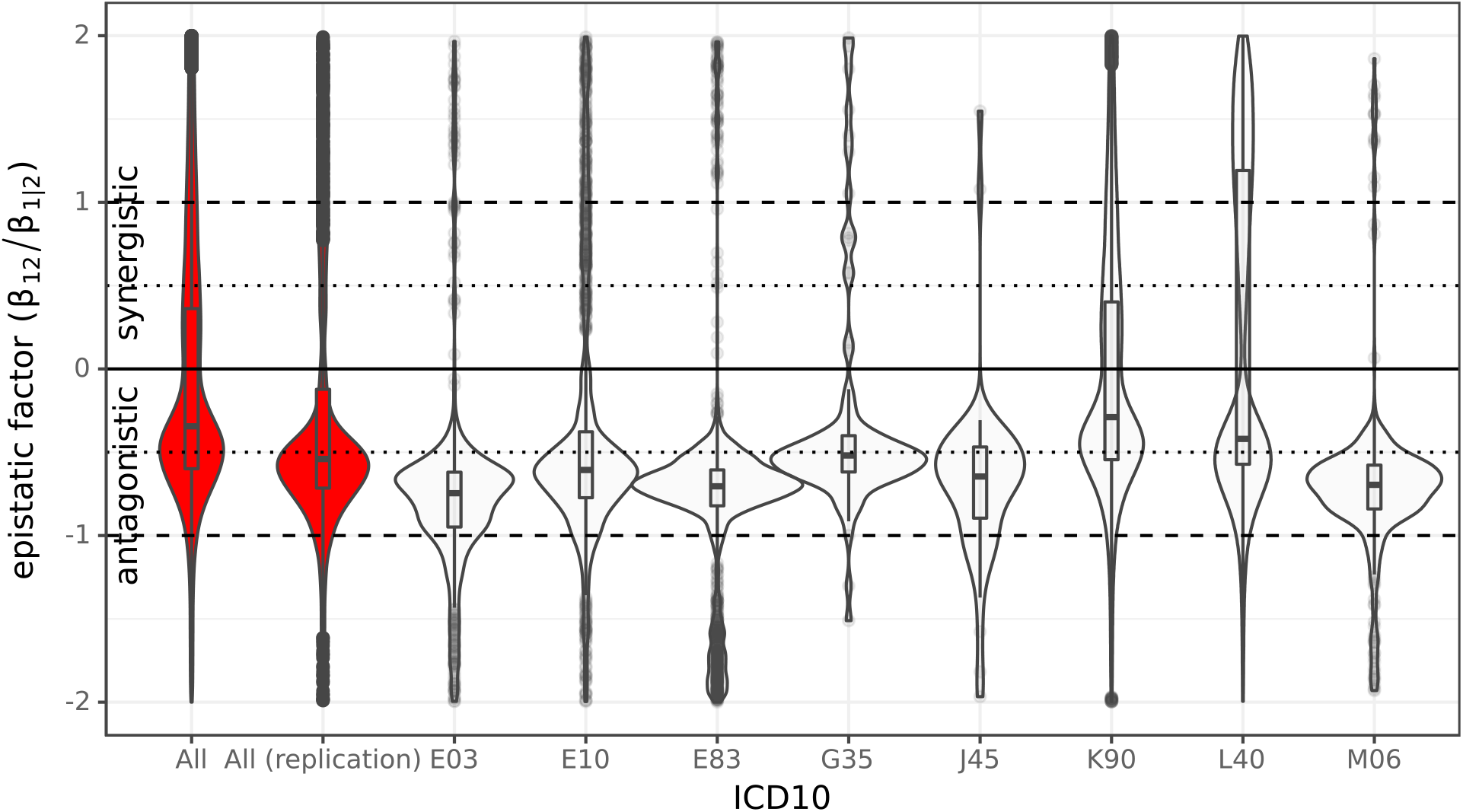
Distribution of epistatic factor β_12_/β_1|2_ derived from the full bivariate logistic model with β_1|2_ = β_1_ (resp. β_1|2_ = β_2_) if only variant 1 (resp. variant 2) presents marginal association (Fisher’s exact test pval < 8.67e-8). If both variants presented marginal association both epistatic factors were considered. Interactions without marginal association were excluded. The category ‘All’ includes all variants pairs significant in the discovery phase and the category ‘All replicated’ all variant pairs replicated.

#### Directionality of epistatic effect

For variants presenting significant marginal association we can derive from the full bivariate logistic model the effect of variant *x*_1_ conditional on variant *x*_2_ as 1 + *β*_12_/*β*_1_*x*_2_ following the approach of Hansen et al. ^44^. The epistatic factor *β*_12_/*β*_1_ quantifies how much the effect of variant *x*_1_ is influenced by variant *x*_2_. Surprisingly we observed that a significant majority (60%) of epistatic interactions were antagonistic (*β*_12_/*β*_1_ < 0). This biais toward antagonistic epistasis is systematically observed for all diseases with more than 100 significant interactions (Figure 7). For E10, E83, G35, E03, M06, and J45, 58% to 79% of interactions with at least one marginal effect presented an antagonistic epistatic factor *β*_12_/*β*_1_ < −0.5. For these interactions the effective effect of variant 1 is in opposite direction in individuals homozygous for minor allele of variant 2 (*x*_2_ = 2) and in homozygotes for major allele (*x*_2_ = 0). Within the replicated interactions with at least one marginal effect 62% were antagonistic.

**Figure 7:**
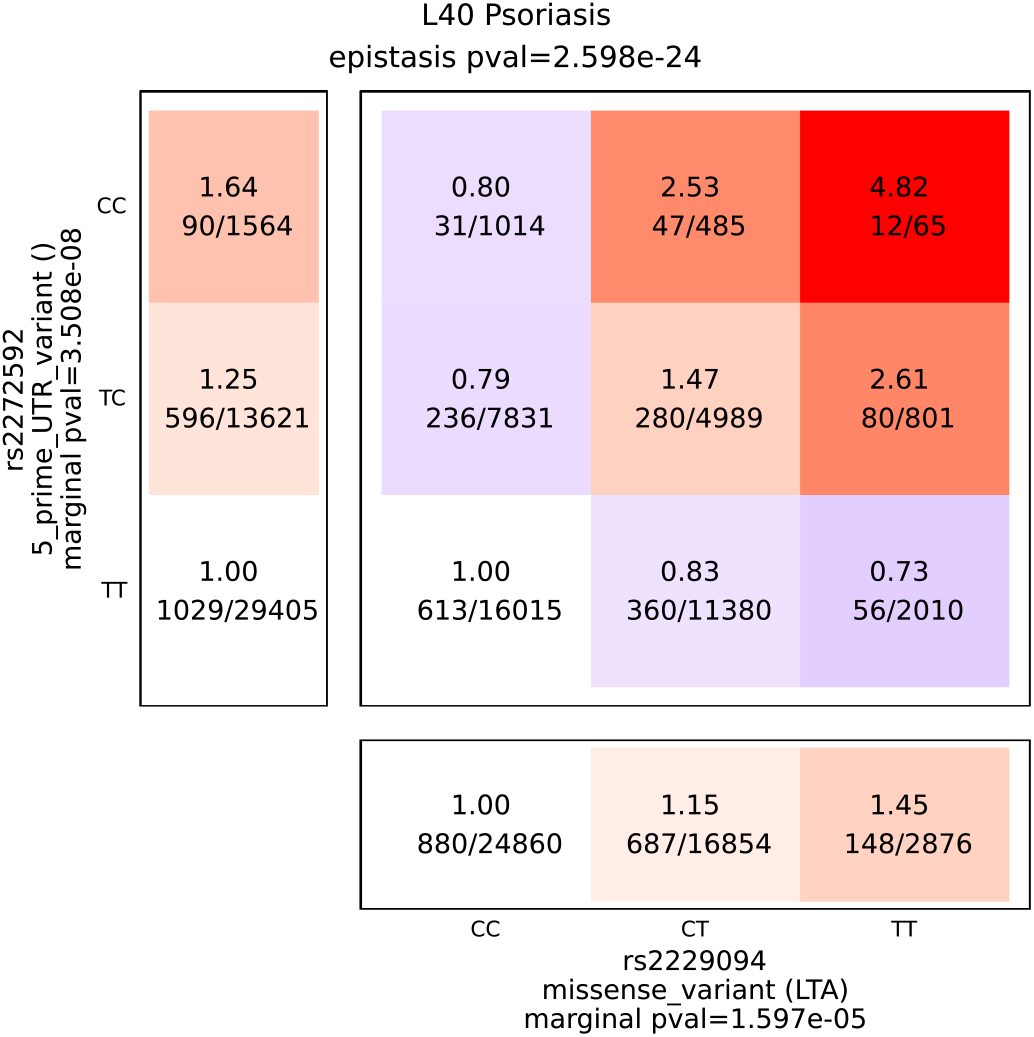
Contingency tables for an antagonistic epistasis in psoriasis involving the LTA missense variant rs3130453. Marginal association pvalue were computed by Fisher’s exact test on the UK biobank psoriasis cohort. Data are represented for the UK Biobank discovery cohort.

#### Examples of antagonistic interactions

Figure 7 presents an example of antagonistic epistasis for psoriasis. Allele C of variant rs2272596 is marginally associated with increased disease risk (OR=1.25 pval=3.5e-8). However the effect of this variant appears to be strongly dependent on variant rs2229094, a LTA missense variant already associated with psoriasis and other immune traits. In presence of rs2229094 common allele disease risk actually decreases with the number of rs2272596-C alleles (OR=0.79) while it strongly increases in presence of rs2229094 minor allele (OR=4.82 for double homozygotes). All antagonistic interactions reproduced in Finngen are reported in Figure S10.

### Variance explained by epistasis

We asked whether the number of associations detected correlated with SNP additive heritability as measured using LD-score regression. Because *h*^*2*^ estimates may be biased downwards by small case counts, we limited the analysis to phenotypes with N_eff_ > 5,000 (N=97). Of those, 22 phenotypes had an estimated *h*^*2*^>0.1, 9 of which displayed at least 1 significant interaction (OR=3.96, Fisher exact test *P*=0.01, Table S1). This enrichment was stronger when considering phenotypes with at least one replicated epistasis (OR=13.2, *P*=0.002), suggesting that diseases with high SNP heritability are also likely to display epistatic effects.

Next, we attempted to estimate the amount of variance explained by epistatic effects. For each variant pairs, with fitted regression models with and without the interaction term, and calculated the difference in pseudo-R2 between both models (Table S3). The mean increase in pseudo-R2 due to the interaction effect was 0.001 (standard deviation=0.001). Therefore, most epistatic events individually did not explain a large fraction of the phenotypic variance. 289 variant pairs displayed a gain in pseudo-R2 > 0.01 when the interaction term was included. Of note, rs1543680 and rs198820 were both additively associated with disorders of mineral metabolism (both P<2e-16), and explained 2.4% of the phenotypic variance. Including the interaction term in the model revealed antagonistic epistasis and together explained 5.6% of the total phenotypic variations. This was true across tested indications, whereas the interactions that explained the most variance were antagonistic (P<2×10^−16^, Figure S9). Therefore, in addition to revealing antagonistic effects, epistasis can explain a good fraction of heritability not captured by additive effects.

### Epistasis mechanism

In the discovery phase 82% of interactions was classified as trans-epistasis, 14% involved variants affecting a same gene and 16% involved variants affecting different genes only (Figure 8). Other interactions would require further functional annotations to identify genes by each variant. Among interactions mapped to different genes a majority (95%) were mapped to different pathways, only 4 (0.02%) were mapped to non HLA genes with known physical interaction, and 891 (4%) to non HLA genes belonging to a same pathway. We observed a similar distribution in all phenotypes with more than 200 interactions. Most replicated interactions were in cis-epistasis in the HLA region (89%), 76% involved variants mapped to the same HLA gene, 16% to different HLA genes. Outside of the HLA region 9 interactions (0.2%) involved variants mapped to the same non HLA genes and 12 (0.3%) to different non HLA genes, 8 belonging to a same pathway.

**Figure 8:**
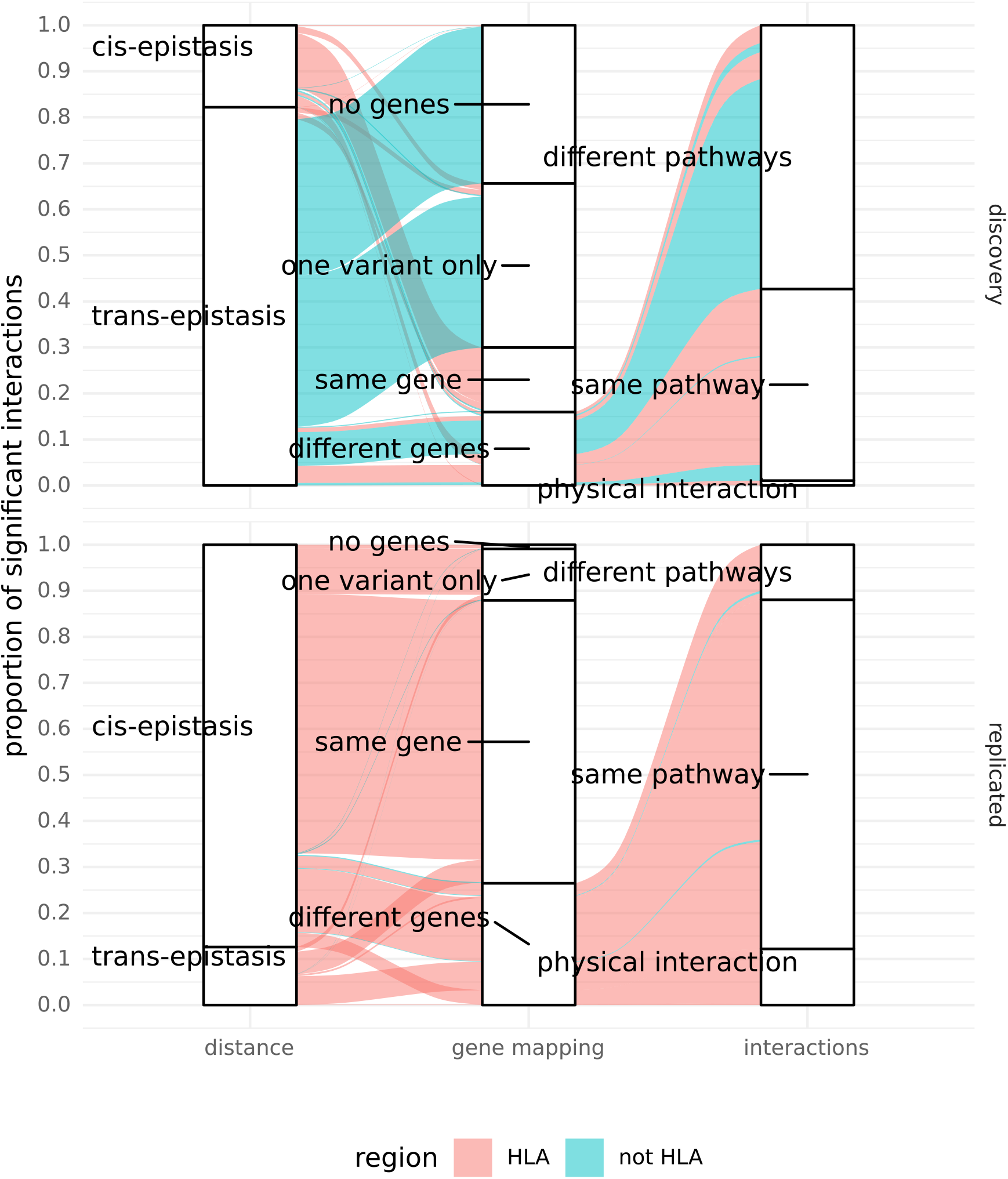
Proportion of cis-epistasis (interaction betwen variants distant of less than 1Mb) and trans-epistasis (left column). Proportion of variant in which both variant were mapped to no gene, only one variant mapped to a gene, both variants mapped to the same gene, and both variants mapped only to different genes (center column). Among interaction mapped to different genes proportion for which both genes are in direct physical interaction, belong to the same canonical pathway or only to different pathways (right column). Alluvium color indicates interactions in HLA region (red) and outside (blue).

## Discussion

In this study we generated a genome-wide map of epistasis across 502 phenotypes. A major goal of this study was to generate this unique resource for genetics researchers interested in investigating epistasis mechanism. The full summary statistics results for all variant pairs with epistasis pval>1e-3 for all 502 phenotypes included in the discovery study are available online (*[URL](https://magellan-edge.sanofi.com/pipeline-64065-3838-1/epistasis/)*). In this article we have explored global properties of epistasis across diseases and provided few illustrative examples of epistasis.

While epistasis has long been considered to be ubiquitous in human genetics it has often been thought to involve small effect sizes requiring prohibitive sample size to reach satisfying statistical power ^25^. This point of view is moderated by our results which show that a moderately powered study is able to identify genome wide significant epistasis in 14% of the phenotypes considered. Previous univariate studies in UK Biobank could identify significant association in a larger proportion of phenotypes, 79% in Canela-Xandri study^4^, but this difference might be explain by the lower statistical power of the present study. Moreover, we show the presence of epistasis between common variants with intermediate to large effect size (OR>2) in 10 diseases. This observation support previous predictions that epistasis can maintain variants with deleterious effect at high frequency over long evolutionary periods ^45^. It also suggests that epistasis would support the hypothesis of common diseases common variants ^46^. The presence of high effect size interaction (OR>5 or <0.2) between intermediate frequency variants (*f* ≈ 0.05) for 3 phenotypes might still suggest the existence of strong effect size epistasis between rare variants. Interestingly all these interactions with high effect size were protective suggesting the presence of strong modulators of genetic disease risk in this space which might give insight into new therapeutic targets for genetically defined disease subgroups. However, exploring epistasis between rare variants would require larger sample sizes or focus studies and we argue that our results show that there is still much to explore in the area of epistasis between common variants.

For most diseases the majority of variants in interactions did not present marginal association. We have shown that even using approaches with more statistical power, such as the pan-UKB analysis with SAIGE approach ^5^, most of these variants (79%) would remain undetected. This observation is consistent with the results of Lippert et al. ^19^ on the 7 diseases of the WTCCC who found interactions between marginally significant SNPs only in T1D. It demonstrates that epistasis can therefore opens the door to new locus discovery. It also adds to existing evidence ^16^ supporting the fact that part of the missing heritability would be explained by the presence of epistasis.

While previous genome-wide epistasis studies on the WTCCC cohorts identified significant interactions almost exclusively in the HLA region ^18,19^ our results suggests the presence of epistasis throughout the genome. However, most interactions replicated in Finngen are in the HLA region. While it is probably due to a stronger association in this region it might suggest that a higher variability of epistasis between populations.

The significant bias towards antagonistic interactions (60% of all interactions in average and more than 95% for several diseases) indicates that epistasis tends to stabilize the phenotype towards population average rather than towards the extremes. Genetic studies in mice have already suggested this mechanism ^47^. Such bias towards antagonistic, or negative, epistasis has been hypothesized to be central in evolution ^48,49^. Negative epistasis has been related to robustness and genetic redundancy in complex organisms^49^ and predicted to decelerate response to selection while contributing to the emergence of population polymorphism by generating multiple equilibria ^50^. Its emergence has been suggested to be a consequence of sexual reproduction ^51^. We show that antagonistic epistatic effect can completely reverse the effect of a variant presenting marginal association and that it is a common phenomenon in human genetics. Epistasis scan therefore opens the way to identify genes with a strong modifying effect on known genetic risk factors. It also shows that epistasis could mask true variant effect and should be taken into account when experimentally validating the effect of genetic variant. The interaction in psoriasis between CCHCR1, MUC22 and HSPA1L variants provides a good illustration. CCHCR1 stop gain rs3130453 is a frequent variant presenting a significant marginal association with moderate increased risk of psoriasis (OR=1.49 pval=7.7e-20). However its effect is small to inexistant (OR=1.2 and OR=1 respectively) in major alleles carriers of MUC22 or HSPA1L common missense variants rs3094672 and rs2227956. An experimental validation of the CCHCR1 variant effect in reference genotypes, by CRISPR editing for instance, would thus be probably unsuccessful. Taking into account epistasis we show that CCHCR1 variant effect is actually intermediate to high (OR=3.2 and 3.1 respectively) in homozygotes minor allele carriers of MUC22 or HSPA1L variants, which could guide experimental validation.

The causal biological mechanism behind statistical epistasis can occur at different levels. In this study we investigated the following mechanisms: variants modifying the structural and transcription properties of a single gene, modifying genes coding for two proteins in direct physical interactions, modifying genes involved in a same pathway, or modifying pathway functions interacting at a higher level. Outside the HLA region we find most variant interacting in trans-epistasis. For a majority of interactions (70%) we did not identify the potential genes affected by the variants, suggesting that the interpretation of epistasis is still limited by our knowledge in functional genomic, at least as represented in the annotation data from open target genetics and ensembl used in this study. Our evaluation, therefore partial, indicates that intra-genes interaction is rare outside the HLA region and that most epistasis occurs at the supra pathway level (94% of mapped non HLA interactions).

We expect that the summary statistics and findings generated in this study will foster further analysis on epistasis. Fine mapping analysis on identified locus in particular would be important to localize the potential causal variants.

## Data Availability

All results summary statistics (p<1e-3) will be freely available.

## Acknowledgements

Data for the discovery analyses were obtained from UK Biobank under an approved data request (Ref: 41906). Authors acknowledge the Finngen consortium for access to data used for replication cohorts.

## References

1. Bycroft, C. et al. The UK Biobank resource with deep phenotyping and genomic data. Nature 562, 203–209 (2018).

2. Finngen. https://www.finngen.fi/en/about (2020).

3. Nagai, A. et al. Overview of the BioBank Japan Project: Study design and profile. J. Epidemiol. 27, S2–S8 (2017).

4. Canela-Xandri, O., Rawlik, K. & Tenesa, A. An atlas of genetic associations in UK Biobank. Nat. Genet. 50, 1593–1599 (2018).

5. Efficiently controlling for case-control imbalance and sample relatedness in large-scale genetic association studies | Nature Genetics. https://www.nature.com/articles/s41588-018-0184-y.

6. Pan-UKB team. https://pan.ukbb.broadinstitute.org. (2020).

7. Sakaue, S. et al. A global atlas of genetic associations of 220 deep phenotypes. medRxiv 2020.10.23.20213652 (2020) doi:10.1101/2020.10.23.20213652.

8. Ge, T., Chen, C.-Y., Neale, B. M., Sabuncu, M. R. & Smoller, J. W. Phenome-wide heritability analysis of the UK Biobank. PLOS Genet. 13, e1006711 (2017).

9. Buniello, A. et al. The NHGRI-EBI GWAS Catalog of published genome-wide association studies, targeted arrays and summary statistics 2019. Nucleic Acids Res. 47, D1005–D1012 (2019).

10. Watanabe, K. et al. A global overview of pleiotropy and genetic architecture in complex traits. Nat. Genet. 51, 1339–1348 (2019).

11. Moore, J. H. The ubiquitous nature of epistasis in determining susceptibility to common human diseases. Hum. Hered. 56, 73–82 (2003).

12. Breen, M. S., Kemena, C., Vlasov, P. K., Notredame, C. & Kondrashov, F. A. Epistasis as the primary factor in molecular evolution. Nature 490, 535–538 (2012).

13. Hemani, G. et al. Detection and replication of epistasis influencing transcription in humans. Nature 508, 249–253 (2014).

14. Lehner, B. Molecular mechanisms of epistasis within and between genes. Trends Genet. 27, 323–331 (2011).

15. Young, A. I. & Durbin, R. Estimation of Epistatic Variance Components and Heritability in Founder Populations and Crosses. Genetics 198, 1405–1416 (2014).

16. Zuk, O., Hechter, E., Sunyaev, S. R. & Lander, E. S. The mystery of missing heritability: Genetic interactions create phantom heritability. 6.

17. Young, A. I. et al. Relatedness disequilibrium regression estimates heritability without environmental bias. Nat. Genet. 50, 1304–1310 (2018).

18. Wan, X. BOOST: A Fast Approach to Detecting Gene-Gene Interactions in Genome-wide Case-Control Studies. 16.

19. Lippert, C. et al. An Exhaustive Epistatic SNP Association Analysis on Expanded Wellcome Trust Data. Sci. Rep. 3, 1099 (2013).

20. Novel methods for epistasis detection in genome-wide association studies. https://journals.plos.org/plosone/article?id=10.1371/journal.pone.0242927.

21. Upton, A., Trelles, O., Cornejo-García, J. A. & Perkins, J. R. Review: High-performance computing to detect epistasis in genome scale data sets. Brief. Bioinform. 17, 368–379 (2016).

22. Niel, C. A survey about methods dedicated to epistasis detection. Front. Genet. 6, 19 (2015).

23. Genome-wide association study of 14,000 cases of seven common diseases and 3,000 shared controls | Nature. https://www.nature.com/articles/nature05911.

24. Goudey, B. et al. GWIS −model-free, fast and exhaustive search for epistatic interactions in case-control GWAS. 18 (2013).

25. Wei, W.-H., Hemani, G. & Haley, C. S. Detecting epistasis in human complex traits. Nat. Rev. Genet. 15, 722–733 (2014).

26. Chatelain, C., Durand, G., Thuillier, V. & Augé, F. Performance of epistasis detection methods in semi-simulated GWAS. BMC Bioinformatics 19, 231 (2018).

27. Luca, D. et al. On the Use of General Control Samples for Genome-wide Association Studies: Genetic Matching Highlights Causal Variants. Am. J. Hum. Genet. 82, 453–463 (2008).

28. FlashPCA2: principal component analysis of Biobank-scale genotype datasets | Bioinformatics | Oxford Academic. https://academic.oup.com/bioinformatics/article/33/17/2776/3798630.

29. Patterson, N., Price, A. L. & Reich, D. Population Structure and Eigenanalysis. PLoS Genet. 2, 20 (2006).

30. Müllner, D. fastcluster: Fast Hierarchical, Agglomerative Clustering Routines for R and Python. J. Stat. Softw. 53, 1–18 (2013).

31. Benjamini, Y. & Hochberg, Y. Controlling the False Discovery Rate: A Practical and Powerful Approach to Multiple Testing. J. R. Stat. Soc. Ser. B Methodol. 57, 289–300 (1995).

32. Ghoussaini, M. et al. Open Targets Genetics: systematic identification of trait-associated genes using large-scale genetics and functional genomics. Nucleic Acids Res. (2020) doi:10.1093/nar/gkaa840.

33. Sun, B. B. et al. Genomic atlas of the human plasma proteome. Nature 558, 73–79 (2018).

34. Mbatchou, J. et al. Computationally efficient whole genome regression for quantitative and binary traits. bioRxiv 2020.06.19.162354 (2020) doi:10.1101/2020.06.19.162354.

35. Bulik-Sullivan, B. K. et al. LD Score regression distinguishes confounding from polygenicity in genome-wide association studies. Nat. Genet. 47, 291–295 (2015).

36. Tervaniemi, M. H. et al. Intracellular signalling pathways and cytoskeletal functions converge on the psoriasis candidate gene CCHCR1 expressed at P-bodies and centrosomes. BMC Genomics 19, (2018).

37. Lin, X., Deng, F.-Y., Lu, X. & Lei, S.-F. Susceptibility Genes for Multiple Sclerosis Identified in a Gene-Based Genome-Wide Association Study. J. Clin. Neurol. Seoul Korea 11, 311–318 (2015).

38. Martine, P. & Rébé, C. Heat Shock Proteins and Inflammasomes. Int. J. Mol. Sci. 20, (2019).

39. Jenkins, S. C., March, R. E., Campbell, R. D. & Milner, C. M. A novel variant of the MHC-linked hsp70, hsp70-hom, is associated with rheumatoid arthritis. Tissue Antigens 56, 38–44 (2000).

40. Boiocchi, C. et al. Heat shock protein 70-hom gene polymorphism and protein expression in multiple sclerosis. J. Neuroimmunol. 298, 189–193 (2016).

41. Takahashi, S. et al. De novo and rare mutations in the HSPA1L heat shock gene associated with inflammatory bowel disease. Genome Med. 9, 8 (2017).

42. Aquino-Gálvez, A. et al. Analysis of heat shock protein 70 gene polymorphisms Mexican patients with idiopathic pulmonary fibrosis. BMC Pulm. Med. 15, (2015).

43. Ambrocio-Ortiz, E. et al. Effect of SNPs in HSP Family Genes, Variation in the mRNA and Intracellular Hsp Levels in COPD Secondary to Tobacco Smoking and Biomass-Burning Smoke. Front. Genet. 10, (2020).

44. Hansen, T. F. & Wagner, G. P. Modeling genetic architecture: a multilinear theory of gene interaction. Theor. Popul. Biol. 59, 61–86 (2001).

45. Hemani, G., Knott, S. & Haley, C. An Evolutionary Perspective on Epistasis and the Missing Heritability. PLOS Genet. 9, 11 (2013).

46. Schork, N. J., Murray, S. S., Frazer, K. A. & Topol, E. J. Common vs. Rare Allele Hypotheses for Complex Diseases. Curr. Opin. Genet. Dev. 19, 212–219 (2009).

47. Tyler, A. L., Donahue, L. R., Churchill, G. A. & Carter, G. W. Weak Epistasis Generally Stabilizes Phenotypes in a Mouse Intercross. PLOS Genet. 12, e1005805 (2016).

48. Barton, N. H. How does epistasis influence the response to selection? Heredity 118, 96–109 (2017).

49. de Visser, J. A. G. M., Cooper, T. F. & Elena, S. F. The causes of epistasis. Proc. R. Soc. B Biol. Sci. 278, 3617–3624 (2011).

50. Hansen, T. F. Why Epistasis Is Important for Selection and Adaptation. Evolution 67, 3501– 3511 (2013).

51. Azevedo, R. B. R., Lohaus, R., Srinivasan, S., Dang, K. K. & Burch, C. L. Sexual reproduction selects for robustness and negative epistasis in artificial gene networks. Nature 440, 87–90 (2006).

